# Atmospheric environment and human brain architecture: associations across multimodal imaging

**DOI:** 10.1101/2025.10.26.25338772

**Authors:** Max Korbmacher, Ole A. Andreassen, Lars T. Westlye, Ivan I. Maximov, Ivan Kuznetsov

## Abstract

Climate change represents one of the most pressing challenges to global public health. Temperature variations, altered precipitation patterns and changes of seasonal and periodic weather events have been linked to a range of adverse health outcomes, including cardiovascular disease, respiratory illness, and mental health disorders. Nevertheless, neurobiological consequences of environmental atmospheric conditions remain poorly characterised in terms of the brain architecture.

We analysed structural neuroimaging data from 30,831 participants in the UK Biobank (4,294 with longitudinal follow-up) and demonstrated that meteorological conditions are associated with measurable variables in brain structure. Specifically, warmer temperatures, greater solar exposure and reduced precipitation and wind speed correlate with differences in both grey matter morphometry and white matter microstructure. Notably, these weather associations exceed in magnitude the contributions of conventional risk factors such as Alzheimer’s disease polygenic risk scores and self-reported mental health status.

These findings expose atmospheric parameters as quantifiable environmental correlates of human cerebral architecture. Thus, the brain tissue may be more sensitive to routine meteorological variation than previously recognised. Due to climate pattern shifts globally, an understanding of these climate-brain interactions becomes increasingly relevant for the biomedical sciences and public health.

## Introduction

Climate change represents a defining challenge to global health system, in particular, the brain health. Rising mean temperatures, shifting precipitation patterns, and increasing frequency of uncommon weather events have been associated with a broad spectrum of adverse health outcomes.^1,2^ Temperature-related mortality in adults over 65 years has increased by 167% compared to the 1990s contributing to approximately 5 million deaths annually worldwide.^3,4^ Beyond mortality, climate variables have been linked to cardiovascular disease, respiratory illness and mental health disorders.^5,6^ Despite the growing evidence on climate-health relationships, the neurobiological consequences of atmospheric conditions remain poorly characterised.

Magnetic resonance imaging (MRI) provides a powerful, non-invasive measures of the human brain structure *in vivo*. MRI contrasts such as T_1_-weighted imaging, enable quantification of grey matter volume and cortical thickness. In turn, diffusion-weighted imaging visualises and provides quantitative metrics of white matter organisation of the human brain. These diffusion metrics serve as sensitive biomarkers of white matter integrity. Large-scale neuroimaging initiatives, most notably the UK Biobank, have acquired standardised MRI data from over 50,000 participants, offering a population-based investigation of brain-environment associations.^7,8^

The simplest evidence linking environmental exposures to brain structure has emerged primarily from air pollution research. Cumulative exposure to fine particulate matter has been associated with reduced white matter volume, with each interquartile increment corresponding to smaller white matter volume^9^. Urban environmental factors, including greenspace proximity, have shown associations with larger brain volumes in UK Biobank analyses, with vitamin D partially mediating these effects as indirect environmental variable.^10^ However, there are no direct investigations of meteorological parameters such as temperature, precipitation, solar radiation and wind affecting brain morphometry and microstructure in the literature.

Multiple biological pathways may link atmospheric conditions to brain structure. Solar radiation influences cutaneous vitamin D synthesis, and vitamin D deficiency has been consistently associated with structural brain changes and elevated dementia risk.^11,12^ Atmospheric conditions modulate circadian rhythms, which, in turn, influence sleeping patterns and perivascular space dynamics implicated in brain waste clearance.^13,14^ Weather also affects physical activity patterns, and exercise-related activity has demonstrated positive associations with brain volumes across multiple regions.^15,16^ Furthermore, thermal stress may influence brain-derived neurotrophic factor regulation, with hot environments attenuating exercise-related its elevations.^17^ Cardiovascular pathways provide additional links, as heart imaging traits display association patterns with brain grey and white matter.^18^

Accumulating evidence from distinct research domains implicates environmental determinants in health outcomes at both the systematic and neurological levels. Current climate projections indicate visible weather pattern changes. Consequently, these changes might affect the brain’s architecture as well, both short- and long-term. In turn, there are many different climate models allowing one to represent different aspects of climate changes and to introduce new weather patterns at these short and long-term. An understanding of neurobiological sensitivity of the brain tissue to atmospheric exposures becomes increasingly relevant and opens a new perspective in neuroscience and neurology. An improved characterisation of weather-brain relationships may inform strategies to anticipate and mitigate climate-related neurological health burdens.

In the presented work, we provide the first systematic investigation of associations between ambient meteorological conditions and brain architecture. Analysing multimodal neuroimaging data from 30,831 participants in the UK Biobank, including 4,294 longitudinal follow-up cases, we examine how temperature, precipitation, solar radiation, and wind speed relate to grey matter morphometry and white matter microstructure.

## Methods and Materials

### Sample

Multimodal MRI data (T_1_-weighted and diffusion-weighted images) were extracted from the UK Biobank database.^7,8^ Participants with mental and behavioural disorders (ICD-10 category F), diseases of the nervous system (ICD-10 category G), diseases of the circulatory system (ICD-10 category I), or stroke were excluded. After merging weather data, polygenic risk scores, and self-reported measures, the final sample comprised N = 30,831 participants (Table 1; follow-up: Supplemental Table 1).

**Table 1.**
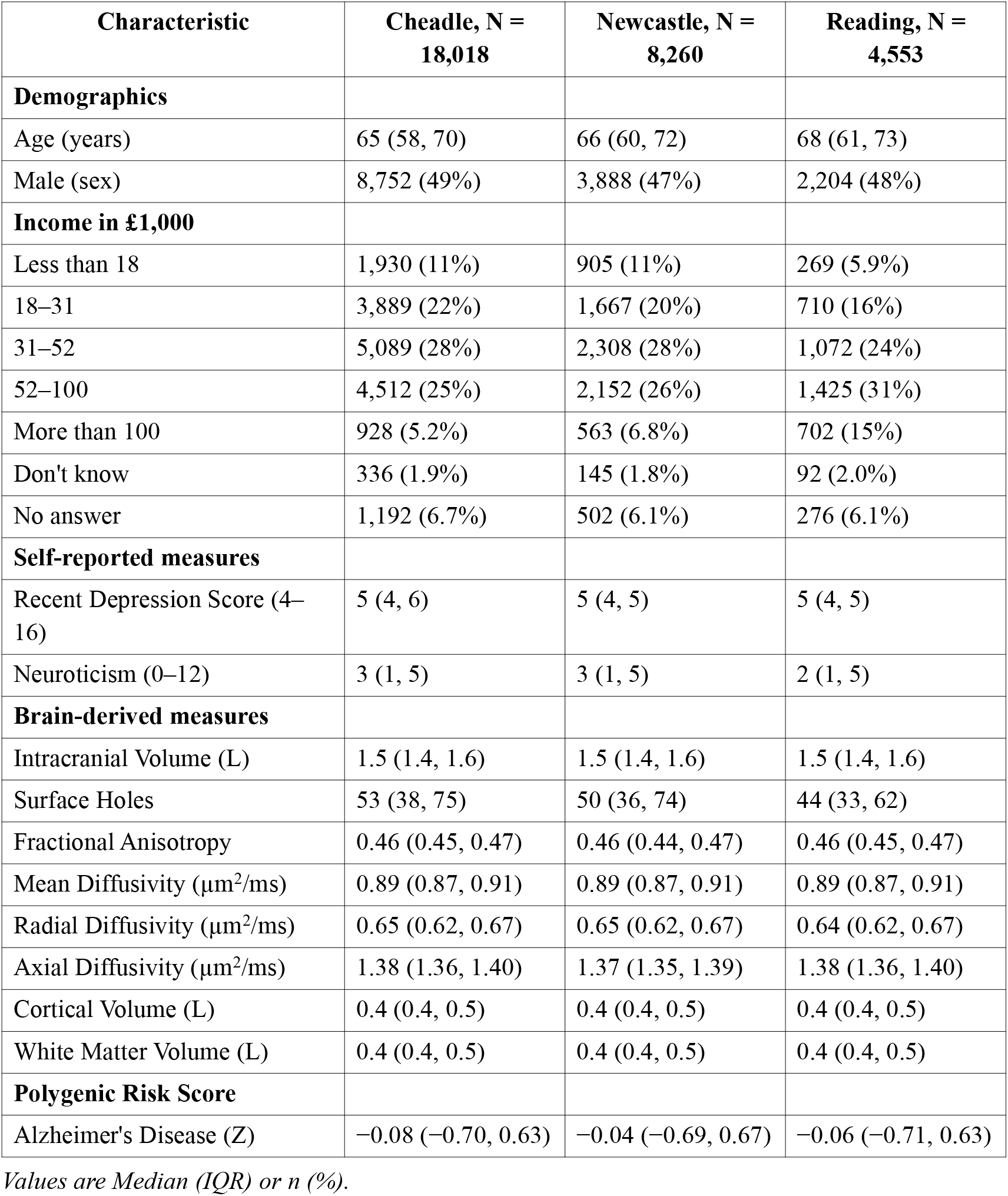
Sample characteristics at baseline.

**Table 2.**
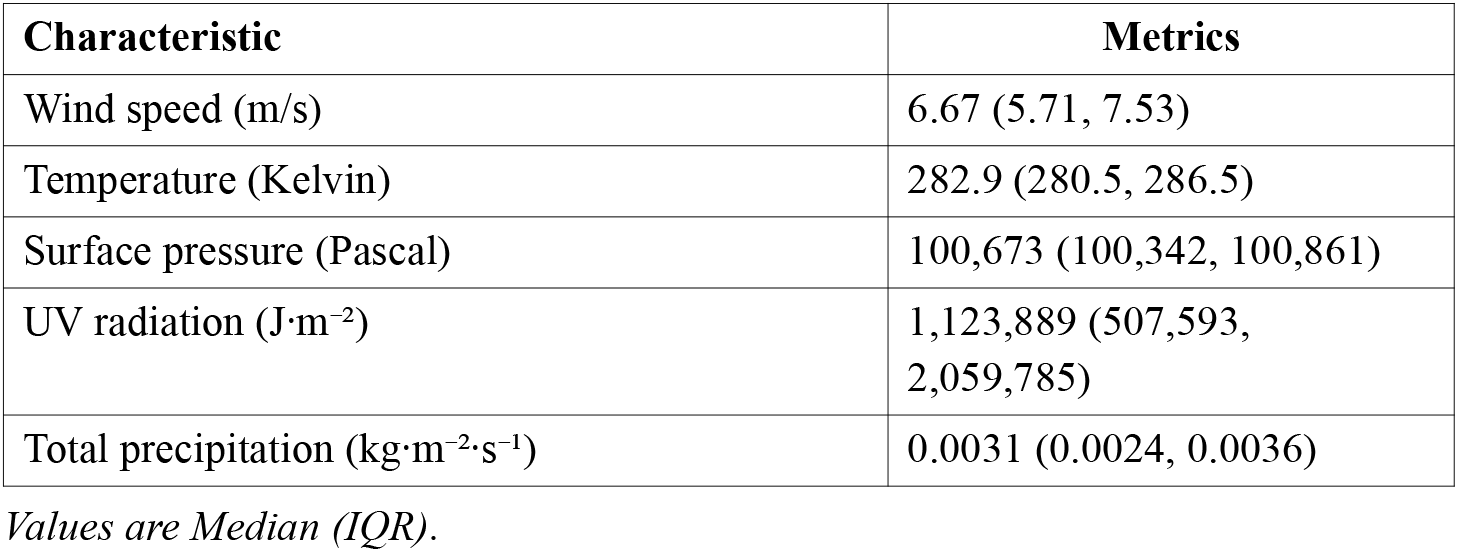
Atmospheric characteristics at baseline.

### MRI Acquisition and Processing

The MRI acquisition protocol has been described previously.^7,8^ Diffusion MRI data were processed using an optimised pipeline^19^ incorporating corrections for noise, Gibbs ringing, susceptibility-induced distortions, motion artefacts, and eddy currents. Isotropic 1 mm^3^ Gaussian smoothing was applied using *FSL* version 6.0.1.^20^ Diffusion tensors^21^ were estimated at each voxel using signal decomposition accounting for kurtosis, yielding more robust estimates.^22^

White matter integrity was analysed using Tract-Based Spatial Statistics.^23^ Briefly, fractional anisotropy images were aligned to MNI space via non-linear registration. A mean fractional anisotropy image and skeleton were generated. Each diffusion parameter map was projected onto this skeleton. For statistical analysis, fractional anisotropy, axial diffusivity, radial diffusivity, and mean diffusivity were averaged across the TBSS derived skeleton. In order to control both an image alignment along the TBSS pipeline and estimated mean diffusion metrics, we employed YTTRIUM,^24^ which converts global diffusion metrics into two-dimensional format using structural similarity extension and excludes non-clustering values.

T_1_-weighted data were processed using *FreeSurfer* version 5.3.0^25^ for surface-based reconstruction and estimation of grey matter, white matter, and intracranial volumes. Surface holes were quantified to avoid bias in large-sample analyses.^26^ T_1_-weighted images with Euler numbers^27^ exceeding three standard deviations from the mean were excluded. Regional parcellation used the Desikan-Killiany Atlas^28^ for T_1_-weighted volumes, yielding 68 regional estimates (34 per hemisphere). White matter was parcellated using the Johns Hopkins University probabilistic atlas^29^ (20 tracts). Each diffusion parameter was averaged across the tracts’ masks, summing to 80 values per individual.

### Polygenic Risk Scores

Polygenic risk scores for Alzheimer’s disease were estimated using LDpred2^30^ with default settings. Summary statistics from a recent genome-wide association study^31^ served as input (minor allele frequency threshold: 0.05). Alzheimer’s disease was selected given well-powered genetic associations and relevance to the ageing sample. Previous work demonstrated associations between late-onset Alzheimer’s disease polygenic risk and brain structure.^32^

### Self-Reported Mental Health Measures

Two self-reported measures were assessed: recent depression and neuroticism.^33^ Depression was quantified using the Recent Depressive Symptoms score (RDS-4; range 4–16), comprising four items covering mood, disinterest, restlessness, and tiredness. Responses were summed across 4-point Likert-type items. Neuroticism was assessed using the Eysenck Personality Questionnaire-Revised Short Form^34^ (12 binary items). Both measures have been validated using UK Biobank test-retest data.^33^

### Atmospheric Data

Monthly meteorological data were obtained from the ERA5 reanalysis dataset,^35^ accessed via the Copernicus Climate Data Store.^36^ ERA5 combines historical observations with numerical weather prediction modelling to produce spatially and temporally complete atmospheric reconstructions spanning over 70 years. Unlike raw observations, reanalysis provides globally gridded fields serving as proxy for actual atmospheric state. This dataset captures long-term climate trends and natural variability.

Spatial means were computed over the United Kingdom (60°–50°N, 7°W–1°E) due to the absence of any information about a birthplace and history of living place for the UK Biobank participants. The larger grid was selected in order to obtain the atmospheric measurements diminishing local variability between recruiting centres. Five metrics were analysed: wind speed at 10 m (m/s), 2-metre temperature (Kelvin), surface downward ultraviolet radiation (J·m^−2^), surface pressure (Pascal), and total precipitation (kg·m^−2^·s^−1^).

### Statistical Analyses

Linear models predicted brain variables (*B*[rain]: cortical thickness, fractional anisotropy, radial diffusivity, axial diffusivity, mean diffusivity) from atmospheric parameters (*W*[eather]) at baseline. Models controlled for site, household income, sex, age, intracranial volume (ICV), and surface holes (SH). ICV influences both diffusion^37^ and volumetric^38^ measures and requires adjustment.

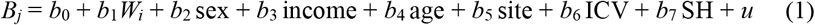

Subsequent models added psychological measures or Alzheimer’s disease polygenic risk scores (PsyG):

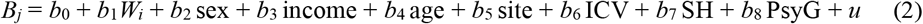

Likelihood ratio tests examined whether psychological and genetic factors improved model performance.

Supplemental analyses included: baseline and longitudinal associations with psychiatric disorder polygenic risk scores using formula 2; linear mixed-effects models with follow-up data and random participant effects; brain age as additional outcome (models trained on cross-sectional data excluding longitudinal participants^39^); and region- and tract-level analyses using Desikan-Killiany and Johns Hopkins University parcellations. Alpha was set at 0.05 with Benjamini-Hochberg correction. Standardised regression coefficients are reported.

## Results

We linked atmospheric data averaged over the United Kingdom for the month preceding health and MRI assessments (Fig.1). Derived atmospheric indices were joined with brain metrics for all 30,831 participants at baseline (Table 1) and for n = 4,294 at follow-up (Supplemental Table 1). As comparison variables, we included self-reported mental health measures and polygenic risk scores (PGRS) for Alzheimer’s disease, given sample age.

**Figure 1.**
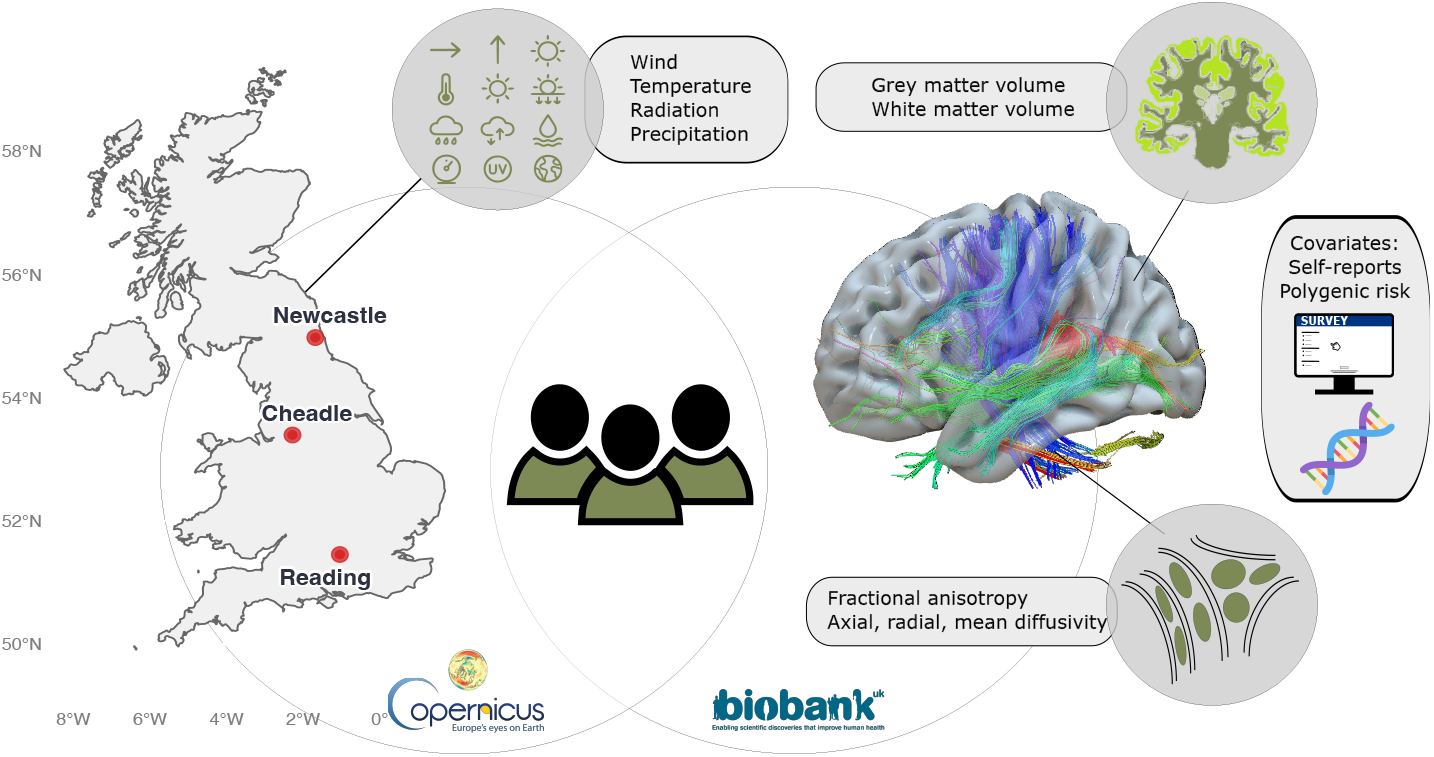
Study design. Monthly meteorological data were linked to structural brain imaging (N = 30,831), yielding measures of grey matter volume, white matter volume, and white matter microstructure (diffusion metrics). Genetic risk scores and self-reported mental health data served as comparison variables.

All analyses controlled for assessment site, household income, sex, age, intracranial volume, and surface holes. Atmospheric variables presented stronger associations with brain structure (median ± MAD |β| = 0.013 ± 0.013) than Alzheimer’s disease PGRS (median ± MAD |β| = 0.006 ± 0.001), and similar magnitude to self-reports of neuroticism and depression (|β| = 0.013 ± 0.012; Fig. 2; Supplemental Data 1). The strongest associations involved fractional anisotropy: 10-meter wind speed (β = −0.059, 95% CI [−0.068, −0.049], p_FDR_ = 1.78 × 10^−30^) and surface downward UV radiation (β = 0.055, 95% CI [0.045, 0.064], p_FDR_ = 4.42 × 10^−27^).

**Figure 2.**
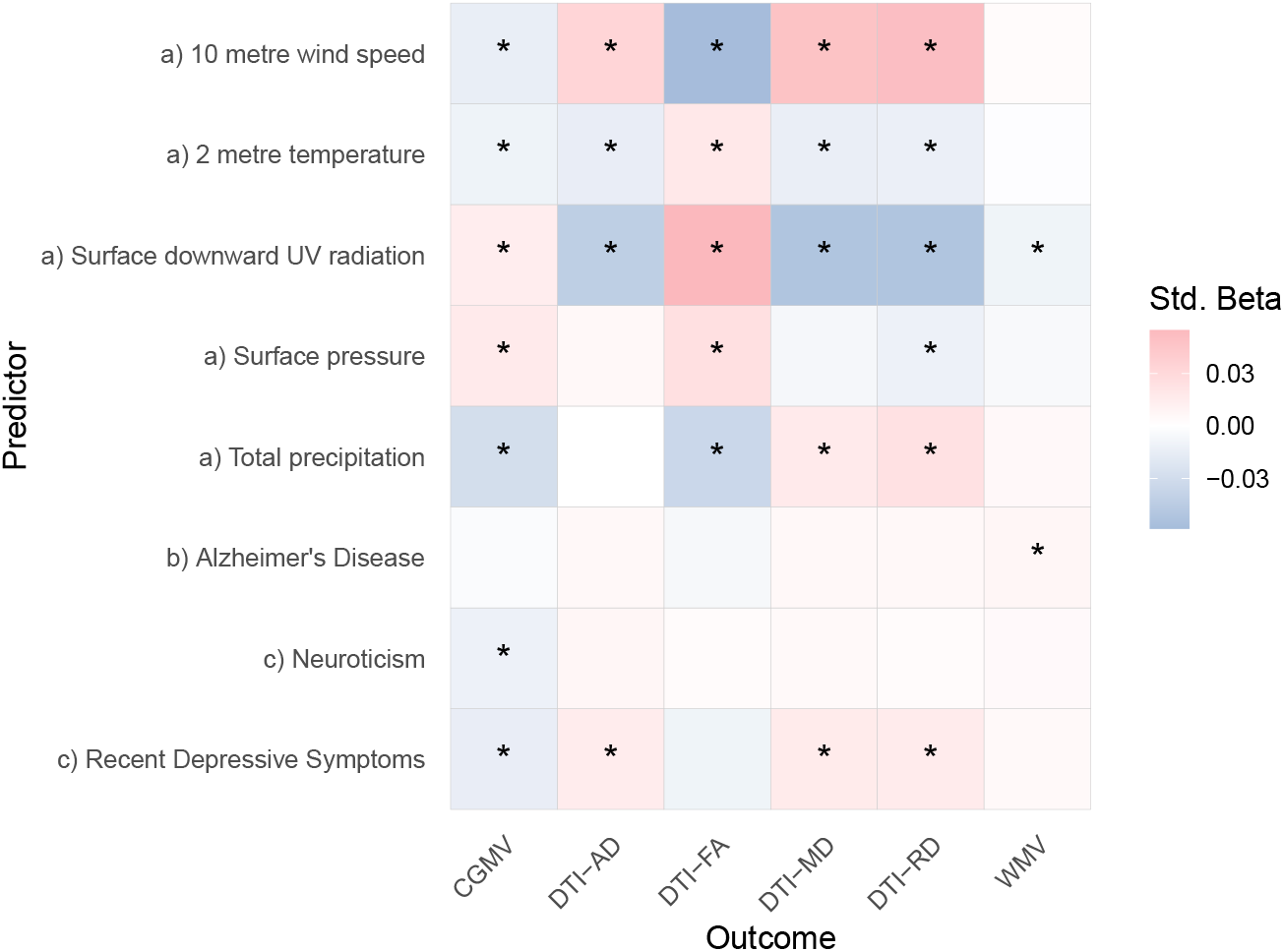
Associations between brain structure and (a) atmospheric indicators, (b) polygenic risk of Alzheimer’s disease, and (c) self-reports of recent depression and neuroticism. PGRS = polygenic risk score; CGMV = cortical grey matter volume; DTI = diffusion tensor imaging; AD = axial diffusivity; FA = fractional anisotropy; MD = mean diffusivity; RD = radial diffusivity; WMV = white matter volume; Std. Beta = standardised regression coefficient.

Favourable weather conditions (lower precipitation, lower wind speed, higher radiation, higher surface pressure) associated with greater cortical volume and higher fractional anisotropy. Fractional anisotropy indicates water diffusion anisotropy along axonal bundles, being a simple proxy for structural integrity of the human brain. Favourable conditions also associated with lower axial, radial, and mean diffusivities, indicating reduced magnitude of diffusivity along and perpendicular to the fibre bundles in the brain (Fig. 2).

Likelihood ratio tests indicated that adding Alzheimer’s disease PGRS or self-reported depression and neuroticism did not improve models predicting brain structure. Changes were non-significant (p > 0.05) in 65 of 90 models (72.2%) after FDR correction. Adding neuroticism to cortical volume models yielded the largest contribution (26.12 < F < 27.16, p < 0.05). However, added variance from PGRS and self-reported outcomes remained low (ΔR^2^ < 0.03%).

The association pattern persisted over time in mixed linear models for n = 4,294 participants with follow-up scans (Figure 3; Supplemental Data 3). Precipitation effects were strongest, associating negatively with cortical volume (β = −0.051, 95% CI [−0.053, −0.048], p_FDR_ < 2.23 × 10^−308^) and fractional anisotropy (β = −0.048, 95% CI [−0.051, −0.046], p_FDR_ < 2.23 × 10^−308^), and positively with mean diffusivity (β = 0.043, 95% CI [0.039, 0.047], p_FDR_ = 2.81 × 10^−108^) and radial diffusivity (β = 0.043, 95% CI [0.039, 0.046], p_FDR_ = 4.93 × 10^−140^).

**Figure 3.**
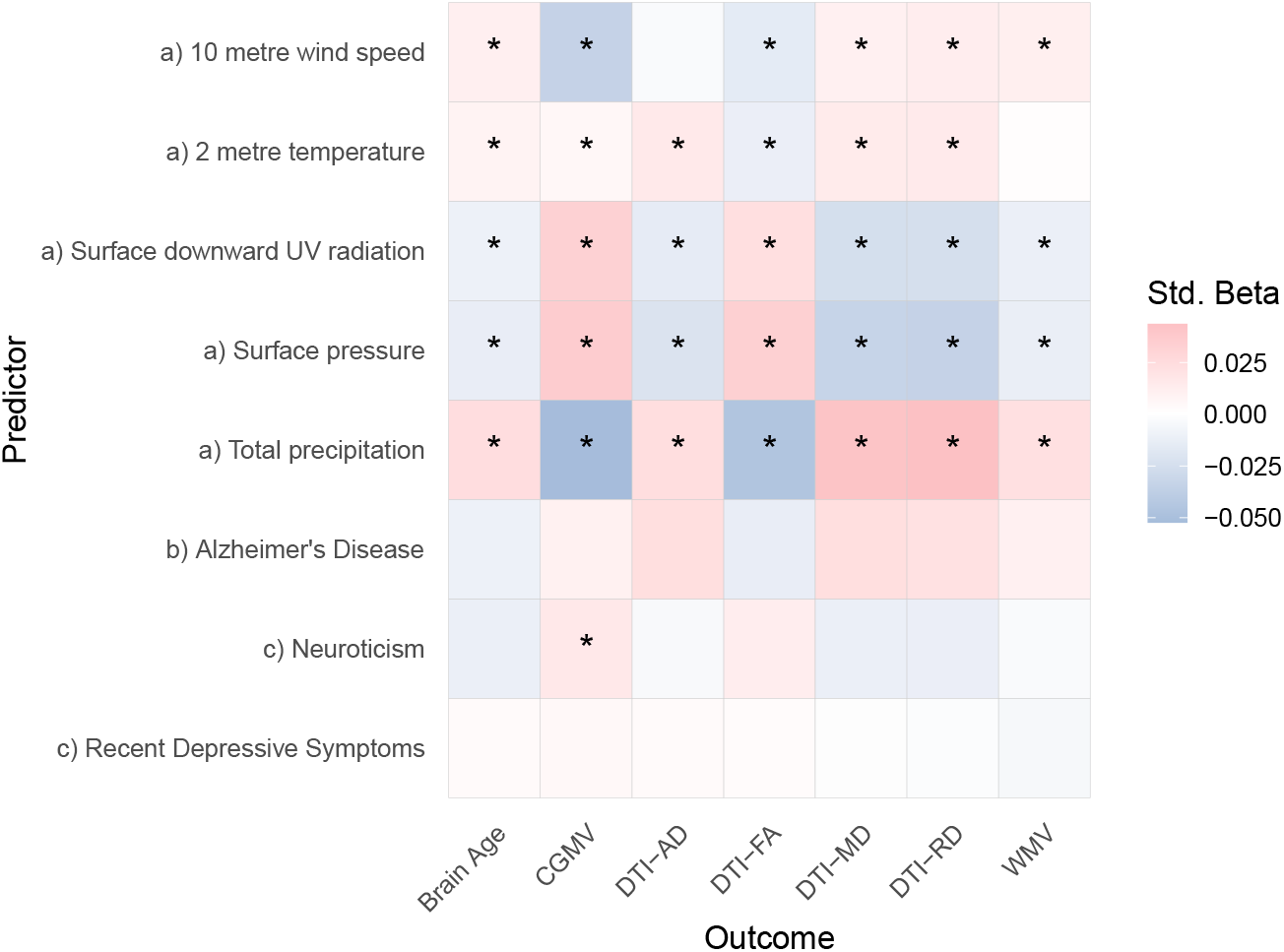
Longitudinal associations between indicators of brain structure and a) weather, b) polygenic risk of Alzheimer’s disease, and c) self-reported outcomes. PGRS = polygenic risk score, CGMV = cortical grey matter volume, DTI = diffusion tensor imaging, AD = axial diffusivity, FA = fractional anisotropy, MD = mean diffusivity, RD = radial diffusivity, WMV = white matter volume. Sample with follow-up: N=4,294.

Given strong cross-sectional and longitudinal associations for precipitation, surface pressure, and wind speed (Fig. 2; Fig. 3), we assessed tract- and region-level brain metrics (Fig 4). White matter microstructure was more sensitive to atmospheric conditions than grey matter volume. Amygdala and ventricular volumes were among the strongest correlates of total precipitation (Fig. 4a). The inferior longitudinal fasciculus presented the strongest associations (β < 0.08) with multiple atmospheric variables across tract-level analyses (Supplemental Data 4).

**Figure 4.**
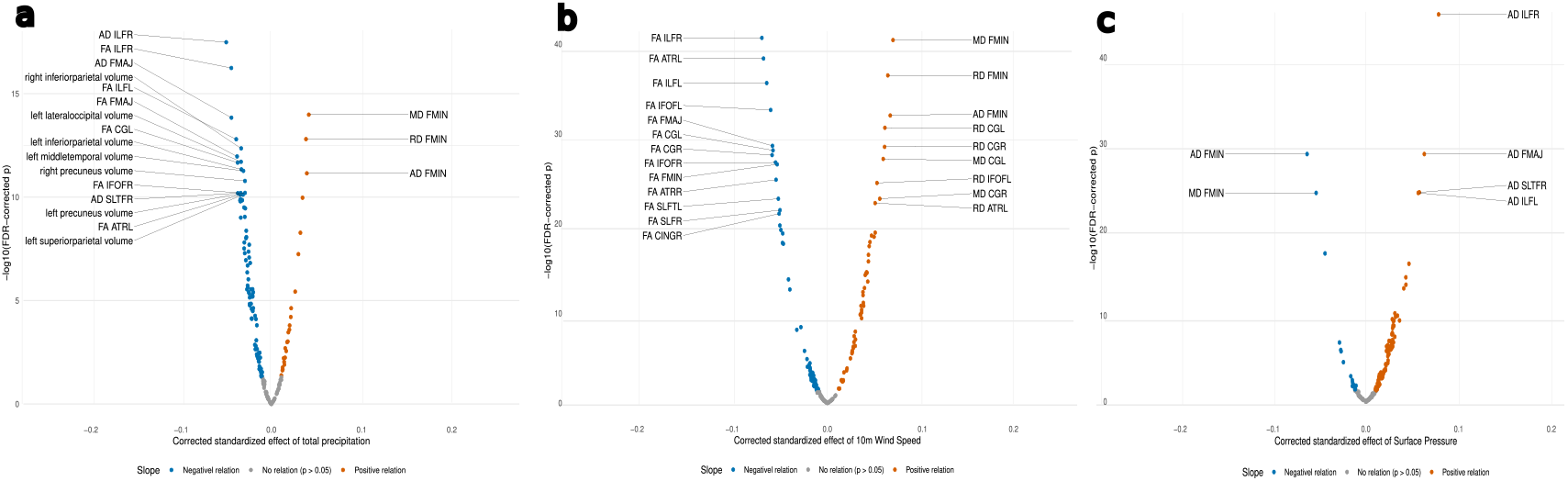
a) Regional and tract-level cross-sectional associations of total precipitation; b) Regional and tract-level cross-sectional associations of wind speed at 10 metres; c) Regional and tract-level cross-sectional associations of surface pressure

## Discussion

In the proposed work we used the most representative measures of brain architecture such as grey morphometry and white matter microstructure and revealed their associations with atmospheric weather metrics. Our findings demonstrated associations of brain architecture with coarse averaged weather parameters. Several biological mechanisms may underlie weather-brain relations. Solar radiation, positively associated with structural integrity, could act through vitamin D synthesis or circadian regulation affecting neuroplasticity and cellular maintenance. Atmospheric pressure and precipitation may influence brain structure indirectly via behavioural pathways. Favourable weather supports outdoor activity, social engagement and exposure to natural environments, are the factors linked to brain health. High-pressure systems associate with low atmospheric dispersion and, hence, air pollution, which confers neurotoxic burden and elevated Alzheimer’s disease risk.

Confounding effects warrant consideration. Despite covariate adjustment, atmospheric patterns and brain structure covary with unmeasured socioeconomic and lifestyle factors. Urbanicity, healthcare access, diet, occupation, and cultural practices all influence brain structure and atmospheric exposure. Individuals with greater resources may mitigate adverse weather through housing, transport, or relocation. It might include a length of holidays, in particular, outside of UK. Observed associations may partly reflect unmeasured interactions between meteorological and social determinants.

Exposure measurement limitations exist. Weather data averaged across the United Kingdom spread an effect of local microclimatic variations and individual exposure differences. Time spent outdoors, indoor conditions, vitamin D supplementation, circadian patterns, and air pollution were not captured. The one-month averaging window may not reflect the most relevant temporal scale. A temporal effect of the weather is also difficult to take into account since only two time points were acquired at UK Biobank study at the moment. The UK Biobank imaging sample comprises middle-aged and older adults in relatively good health, limiting generalisability.

Non-linearity and boundary effects may also exist. Extreme weather or cumulative long-term exposure may have distinct impacts from monthly means. Seasonal cycles could exert neurobiological influences distinct from stochastic weather variation. However, previous multi-scan case studies suggest brain structure estimates are relatively robust to seasonal effects. Individual differences in weather sensitivity, shaped by genetic, developmental or health factors, were not examined but may modulate these relationships.

The revealed findings have implications for neuroscience and public health. Longitudinal designs are needed to establish temporal precedence, assess reversibility, and quantify dose-response relationships. Such studies could clarify whether within-person weather exposure variation predicts brain structural change and identify sensitive exposure windows. Future research should examine regional specificity, vulnerable populations, and modifiers including outdoor time, activity levels, and baseline health. Wearable sensor data would improve exposure assessment beyond aggregated meteorological indices.

If causal relationships are confirmed, findings could inform interventions. Urban planning and building design might incorporate strategies to mitigate negative weather effects on neurological health. Individual-level interventions, for example, light therapy, vitamin D supplementation, particular activity during negatively affecting weather conditions, might be included as well. As global climate change alters weather variability and extremes, understanding neurobiological systems sensitive to meteorological variation becomes critical for anticipating future health burdens. In summary, ambient weather patterns associate with the human brain architecture, with effect sizes exceeding those of PGRS for common brain disorders and self-reported mental health measures. These data provide an empirical foundation for environmental neuroscience linking climate variability to brain biology. While causal mechanisms remain to be established, results suggest meteorological conditions are neurobiologically relevant and should be integrated into models of brain health as global environmental conditions shift.

## Data Availability

Materials and code are available at https://github.com/MaxKorbmacher/WeatherBrain

## Notes

### Competing Interest Statement

MK has received a speaker's honorarium from Merck.
OAA has received a speaker's honorarium from Lundbeck, Janssen, Otsuka and Lilly, and is a consultant to Coretechs.ai and Precision Health.
LTW is a minor shareholder of baba.vision.

### Funding Statement

No non-public funds were used for this study.
Please see the pdf for the full funding statement.

### Author Declarations

The UK Research Ethics Committee (REC) approval reference for UK Biobank is 16/NW/0274.

### Summary of Updates

Added figures from supplement, re-worked writing: sharpened, shortened, re-phrasing.

