## Supplement for "Atmospheric environment and human brain architecture: associations across multimodal imaging"

### Supplemental Information

Supplement for the article “Atmospheric environment and human brain architecture: associations across multimodal imaging” by Max Korbmacher et al. (2025).

#### Supplemental Tables

**Supplemental Table 1. Sample characteristics at follow-up.**

| Characteristic | Cheadle, N = 2,424[1] | Newcastle, N = 1,674[1] | Reading, N = 196 |
|---|---|---|---|
| *Demographics* | | | |
| Age (years) | 64 (58, 70) | 65 (59, 70) | 65 (60, 72) |
| Male (sex) | 1,146 (47%) | 768 (46%) | 82 (42%) |
| *Income in £1,000* | | | |
| Less than 18 | 218 (9.0%) | 154 (9.3%) | 14 (7.2%) |
| 18-31 | 598 (25%) | 397 (24%) | 31 (16%) |
| 31-52 | 706 (29%) | 461 (28%) | 51 (26%) |
| 52-100 | 529 (22%) | 397 (24%) | 65 (33%) |
| More than 100 | 171 (7.1%) | 104 (6.3%) | 24 (12%) |
| Don't know | 61 (2.5%) | 44 (2.7%) | 5 (2.6%) |
| No answer | 132 (5.5%) | 101 (6.1%) | 5 (2.6%) |
| *Self-reported measures* | | | |
| Recent Depression Score (range: 4-16) | 4 (4, 5) | 4 (4, 6) | 5 (4, 6) |
| Neuroticism (range: 0-12) | 2 (0, 5) | 2 (0, 5) | 2 (1, 4) |
| *Brain-derived measures* | | | |
| Intracranial Volume (liters) | 1.5 (1,4, 1.6) | 1.5 (1.4, 1.6) | 1.5 (1.4, 1.6) |
| SurfaceHoles | 41 (31, 55) | 41 (31, 55) | 38 (28, 53) |
| Fractional Anisotropy | 0.46 (0.45, 0.47) | 0.46 (0.45, 0.47) | 0.46 (0.45, 0.48) |
| Mean Diffusivity | 0.89 (0.87, 0.92) | 0.88 (0.86, 0.90) | 0.88 (0.87, 0.90) |
| Radial Diffusivity | 0.65 (0.62, 0.67) | 0.64 (0.62, 0.66) | 0.64 (0.62, 0.65) |
| Axial Diffusivity | 1.38 (1.36, 1.40) | 1.37 (1.35, 1.38) | 1.38 (1.36, 1.39) |
| Cortical Volume (liters) | 0.4 (0.4, 0.5) | 0.4 (0.4, 0.5) | 0.4 (0.4, 0.5) |
| White Matter Volume (liters) | 0.5 (0.4, 0.5) | 0.5 (0.4, 0.5) | 0.5 (0.4, 0.5) |
| *Polygenic Risk Scores (Z-scores)* | | | |
| Anxiety Disorder | -0.05 (-0.65, 0.71) | -0.01 (-0.68, 0.67) | -0.18 (-0.95, 0.49) |
| Attention Deficit/Hyperactivity Disorder | 0.02 (-0.73, 0.65) | 0.06 (-0.66, 0.67) | 0.09 (-0.70, 0.79) |
| Autism Spectrum Disorder | -0.05 (-0.69, 0.71) | -0.05 (-0.75, 0.68) | 0.24 (-0.54, 0.73) |
| Bipolar Depressive Disorder | -0.04 (-0.72, 0.67) | 0.06 (-0.59, 0.72) | 0.03 (-0.61, 0.67) |

| | | | |
|---|---|---|---|
| Major Depressive Disorder | 0.01 (-0.65, 0.68) | -0.01 (-0.74, 0.64) | 0.01 (-0.63, 0.62) |
| Obsessive-Compulsive Disorder | 0.01 (-0.68, 0.67) | 0.03 (-0.72, 0.69) | 0.06 (-0.51, 0.69) |
| Schizophrenia | 0.04 (-0.65, 0.70) | -0.03 (-0.59, 0.66) | -0.16 (-0.85, 0.47) |
| Alzheimer's Disease | -0.04 (-0.67, 0.67) | -0.07 (-0.69, 0.59) | -0.04 (-0.60, 0.57) |

[1] Median (IQR); n (%)

**Supplemental Table 2. Baseline sample characteristics including polygenic risk scores of psychiatric disorders.**

| Characteristic | Cheadle, N = 18,018[1] | Newcastle, N = 8,260[1] | Reading, N = 4,553[1] |
|---|---|---|---|
| *Demographics* | | | |
| Age (years) | 65 (58, 70) | 66 (60, 72) | 68 (61, 73) |
| Male (sex) | 8,752 (49%) | 3,888 (47%) | 2,204 (48%) |
| *Income in £1,000* | | | |
| Less than 18 | 1,930 (11%) | 905 (11%) | 269 (5.9%) |
| 18-31 | 3,889 (22%) | 1,667 (20%) | 710 (16%) |
| 31-52 | 5,089 (28%) | 2,308 (28%) | 1,072 (24%) |
| 52-100 | 4,512 (25%) | 2,152 (26%) | 1,425 (31%) |
| More than 100 | 928 (5.2%) | 563 (6.8%) | 702 (15%) |
| Don't know | 336 (1.9%) | 145 (1.8%) | 92 (2.0%) |
| No answer | 1,192 (6.7%) | 502 (6.1%) | 276 (6.1%) |
| *Self-reported measures* | | | |
| Recent Depression Score (range: 4-16) | 5 (4, 6) | 5 (4, 5) | 5 (4, 5) |
| Neuroticism (range: 0-12) | 3 (1, 5) | 3 (1, 5) | 2 (1, 5) |
| *Brain-derived measures* | | | |
| Intracranial Volume (liters) | 1.5 (1,4, 1.6) | 1.5 (1.4, 1.6) | 1.5 (1.4, 1.6) |
| SurfaceHoles | 53 (38, 75) | 50 (36, 74) | 44 (33, 62) |
| Fractional Anisotropy | 0.46 (0.45, 0.47) | 0.46 (0.44, 0.47) | 0.46 (0.45, 0.47) |
| Mean Diffusivity | 0.89 (0.87, 0.91) | 0.89 (0.87, 0.91) | 0.89 (0.87, 0.91) |
| Radial Diffusivity | 0.65 (0.62, 0.67) | 0.65 (0.62, 0.67) | 0.64 (0.62, 0.67) |
| Axial Diffusivity | 1.38 (1.36, 1.40) | 1.37 (1.35, 1.39) | 1.38 (1.36, 1.40) |
| Cortical Volume (liters) | 0.4 (0.4, 0.5) | 0.4 (0.4, 0.5) | 0.4 (0.4, 0.5) |
| White Matter Volume (liters) | 0.4 (0.4, 0.5) | 0.4 (0.4, 0.5) | 0.4 (0.4, 0.5) |
| *Polygenic Risk Scores (Z-scores)* | | | |
| Anxiety Disorder | -0.01 (-0.66, 0.67) | -0.01 (-0.68, 0.66) | -0.05 (-0.71, 0.62) |
| Attention Deficit/Hyperactivity Disorder | 0.01 (-0.66, 0.68) | 0.02 (-0.66, 0.68) | -0.06 (-0.74, 0.63) |
| Autism Spectrum Disorder | 0.01 (-0.67, 0.68) | -0.02 (-0.69, 0.65) | 0.02 (-0.65, 0.68) |
| Bipolar Depressive Disorder | -0.02 (-0.69, 0.66) | 0.03 (-0.62, 0.71) | 0.04 (-0.65, 0.68) |

| | | | |
|---|---|---|---|
| Major Depressive Disorder | 0.00 (-0.66, 0.68) | -0.02 (-0.69, 0.64) | -0.02 (-0.71, 0.67) |
| Obsessive-Compulsive Disorder | 0.01 (-0.66, 0.68) | -0.04 (-0.73, 0.63) | 0.04 (-0.66, 0.73) |
| Schizophrenia | 0.05 (-0.65, 0.70) | 0.02 (-0.65, 0.69) | -0.03 (-0.71, 0.66) |
| Alzheimer's Disease | -0.08 (-0.70, 0.63) | -0.04 (-0.69, 0.67) | -0.06 (-0.71, 0.63) |

[1] Median (IQR); n (%)

#### Supplemental Notes

##### Supplemental Note 1. Supplemental Methods: Polygenic risk scores of psychiatric disorders

For additional investigation of polygenic risk scores (PGRS), we estimated PGRS, as in the main text, for each participant with available genomic data, using LDPred2[1] with default settings. As input for the PGRS, we used summary statistics from recent genome-wide association studies of Autism Spectrum Disorder (ASD),[2] Major Depressive Disorder (MDD)[3], Schizophrenia (SCZ),[4] Attention Deficit Hyperactivity Disorder (ADHD),[5] Bipolar Disorder (BIP),[6] Obsessive Compulsive Disorder (OCD),[7] and Anxiety Disorder (ANX).[8] We used a minor allele frequency of 0.05, as the threshold most commonly used in PGRS studies of psychiatric disorders.

#### Supplemental Data Files

**Supplemental Data 1. Coefficients for cross-sectional analyses**

**Supplemental Data 2. Likelihood ratio tests for cross-sectional analyses**

**Supplemental Data 3. Longitudinal analyses**

**Supplemental Data 4. Cross-sectional regional analyses**
